# Intensity of home-based telework and work engagement during the COVID-19 pandemic

**DOI:** 10.1101/2021.04.01.21254795

**Authors:** Tomohisa Nagata, Masako Nagata, Kazunori Ikegami, Ayako Hino, Seiichiro Tateishi, Mayumi Tsuji, Shinya Matsuda, Yoshihisa Fujino, Koji Mori, for the CORoNaWork project

## Abstract

**Objective:** The present study examined the relationship between the intensity of home-based telework and work engagement.

**Methods:** This cross-sectional study using a self-administrated questionnaire survey was conducted from December 22 to 25, 2020, in Japan. The subjects were asked single-item questions about the intensity of telework and three-item questions about work engagement using the Utrecht Work Engagement Scale. Coefficients were estimated using a multilevel regression model nested by prefecture of residence and adjusted for covariates.

**Results:** High-intensity (four or more days per week) telework was not associated with high work engagement for men or women. In contrast, low and moderate intensity (three days per week to once per month) were associated with high work engagement. The results were consistent when stratified by sex.

**Conclusions:** Reasonable-intensity telework may have beneficial effects on work engagement.

**Clinical Significance:** This study revealed that a reasonable intensity of telework may have beneficial effects on work engagement. A reasonable intensity is defined as low (once per week to once per month) or moderate intensity (two to three days per week) for both men and women.

## Introduction

In response to the economic and social strain induced by the COVID-19 pandemic, the Japanese government officially declared a state of emergency on April 8, 2020, in an effort to prevent the collapse of medical services.^1^ While the declaration was initially limited to seven prefectures, it was expanded to include the entire country on April 16, 2020.^2^ To limit the spread of COVID-19, many companies have made major changes to employees’ working style, and the frequency of opportunities to engage in home-based telework has dramatically increased. Indeed, in a November 2020 survey of approximately 20,000 people, the national average telework implementation rate among full-time employees in Japan was 24.7%.^3^

A review paper on the mental health effects of home-based telework showed that telework has increased isolation, depression, stress, and overwork. However, this result is inconsistent with the findings of previous studies.^4^ One study found that employees who telecommuted eight or fewer hours per month were significantly less likely than non-telecommuters to experience depression.^5^ As background to these results, the authors pointed out that the characteristics and conditions of telework (workplace support, autonomy, etc.) are more important than whether one is simply a teleworker or not.^6^ Consideration should be given to not only the negative influences on mental health but also the positive influences of this approach to work. Work engagement, one such positive influence, is a concept characterized by vigor, dedication, and absorption.^7^ In a cross-sectional study describing the relationship between the extent of telecommuting and work engagement, the working environment, such as social support from colleagues, was found to influence work engagement, although no direct influence of telecommuting on engagement was noted.^6^

Many workers were forced into telework without any preparation time due to the relatively sudden appearance of the COVID-19 pandemic.^8,9^ Before the pandemic, telework was often available to individual employees as an option^10^, with a high degree of flexibility afforded them in terms of where and when they could work. However, the working environment and conditions during the pandemic are markedly different from the norm, as workers are often deprived of the choice to telework or not. The question of how home-based telework affects work engagement in this situation is unclear.

The present study examined the relationship between the intensity of home-based telework and work engagement adjusted for work environment, such as workplace support and decision latitude. We also analyzed the results by sex, an approach adopted by many previous studies^11^, as men and women often perform different roles in the household, which is likely to affect the results. One study found less fatigue and stress in men who regularly worked from home than in those who did not; in contrast, less stress but greater fatigue was noted in women who worked from home than in those who did not.^12^ Telework was also shown to be associated with greater stress as well as happiness in male workers, although no such effect was found in female workers.^13^ To our knowledge, however, no studies have examined the relationship between telework and work engagement by sex.

## Methods

A research group from the University of Occupational and Environmental Health, Japan, conducted a prospective cohort study, known as the Collaborative Online Research on Novel-coronavirus and Work study (CORoNaWork study), as a self-administrated questionnaire survey through the internet survey company Cross Marketing Inc. (Tokyo, Japan). During the baseline survey, conducted December 22-25, 2020, Japan was in the midst of its third wave of the pandemic, at which point the number of COVID-19 infections and deaths was markedly higher than in the first and second waves; the country was thus on high alert.

A portion of the baseline survey from the CORoNaWork study was used to conduct the present cross sectional study. The study protocol, including the sampling plan and subject recruitment procedure, has been previously reported in detail.^14^ Participants were aged 20-65 years and working at the time of the baseline survey (n = 33,087 total). Participants in the CORoNaWork study were stratified by cluster sampling according to gender, age, and region. After excluding 6,051 initial subjects who provided invalid responses, we ultimately included 27,036 in the database. We analyzed the 19,659 workers remaining after further excluding self-employed workers (2,709), workers in small/home offices (2,721) and agriculture, forestry, and fishing workers (1,947) to meet the research purposes.

The present study was approved by the Ethics Committee of the University of Occupational and Environmental Health, Japan.

### Measures

#### Assessment of Intensity of Home-Based Telework

We asked subjects, “Do you work at home? Please choose the answer that is closest to your current situation”, and respondents chose one of the following five options: Four days a week or more; Two to three days a week; One day a week; More than once a month but less than once a week; and Never. Participants were divided into four groups by intensity of telework: high intensity for telework ≥4 days/week, moderate intensity for telework 2-3 days/week, low intensity for telework once a week to once a month, and no telework for those without teleworking.

#### Assessment of Work Engagement

The three-item Japanese version of the Utrecht Work Engagement Scale (UWES-3) was used to assess work engagement.^15,16^ The UWES-9 has previously been translated into Japanese, and the Japanese version was found to have acceptable internal consistency and reliability as well as factor and construct validity.^15^ The items of UWES-3 were selected from among those included in the UWES-9. The UWES-3 has been validated in five countries, including Japan^16^ and includes measures of vigor (one item), dedication (one item), and absorption (one item), with each item measured on a seven-point response scale ranging from 0 (never) to 6 (always/every day). Overall scores on the UWES-3 (range: 0 to 6) are calculated by averaging the individual item scores. Cronbach’s α coefficient for the total UWES-3 score in this study was 0.92.

#### Assessment of Covariates

Covariates included demographic, socioeconomic factors, occupation and industry, psychological demands, decision latitude, and workplace support. Age was expressed as a continuous variable. Education was classified into five categories: junior high school, high school, junior college or technical school, university, and graduate school. Yearly household income was classified into four categories: <2.50 million Japanese yen (JPY); 2.50-3.75 million JPY; 3.75-5.25 million JPY; and >5.25 million JPY. Marital status was classified into three categories: married; divorced or widowed; and never married. Presence of family living together was classified into two categories: present and absent. In this survey, participants chose 1 of 10 options for their occupation: general employee; manager; executive manager; public employee, faculty member, or non-profit organization employee; temporary/contract employee; self-employed; small office/home office (SOHO); agriculture, forestry, or fishing; professional occupations (lawyer, tax accountant, medical-related, etc.); and other occupations. Three of these categories were excluded from this study, as mentioned above, so the occupations were ultimately classified into seven categories. The participants chose 1 of 22 options for their working industry: energy, materials, industrial machinery; food; beverages/tobacco products; pharmaceuticals/medical supplies; cosmetics/toiletries/sanitary products; fashion and accessories; precision machinery and office supplies; home appliances/audio visual equipment; automobiles and transportation equipment; household goods; hobby/sporting goods; real estate and housing equipment; information and communication; distribution and retail; finance/insurance; transportation and leisure; restaurant and other services; public offices and organizations; education, medical services, religion; mass media; market research; and others.

Work-related stress was assessed by the 22-item Japanese version of the Job Content Questionnaire.^17,18^ The Job Content Questionnaire comprises a five-item psychological demands scale (response range 12–48), a nine-item decision latitude scale (response range 24–96), and an eight-item workplace support scale (response range 8–32) created by summing supervisor support and co-worker support. Each item was measured on a four-point Likert type scale ranging from 1 (strongly disagree) to 4 (strongly agree). In previous studies concerning the relationship between telework and work engagement, a theoretical model with three factors mediating the relationship between the two was developed (6). In the present study with the same target population, we also found that telework was related to three factors: psychological demands, decision latitude, and workplace support.^19^

In addition, the prefecture of residence was used as a community-level variable.

### Statistical Analyses

Multilevel regression analyses were used to examine the association between intensity of telework and work engagement. The coefficients and standard errors were estimated using multilevel regression analyses nested by prefecture of residence and adjusted for sex and age (Model 1). We then additionally adjusted for income, marriage, and presence of family living together (Model 2), along with occupation and industry (Model 3), and psychological demand, decision latitude and workplace support (Model 4). We did not adjust for education, as adjusting for education would constitute over-adjustment. In addition, we performed sex-stratified analyses in the same manner.

The level of significance was set at 0.05 (two-tailed). A trend test was conducted by treating the surveyed telework as a continuous variable on a five-point scale (one to five points) in order of decreasing frequency and performing the analysis in the same manner. All analyses were performed using Stata 16SE (StataCorp LLC, College Station, TX, USA).

## Results

The mean age was higher for men than for women. Men also had a higher educational attainment and yearly household income than women. The percentage of married people was higher among men than women. Psychological demands were higher in women, whereas decision latitude was higher in men. No marked difference in workplace support by sex was noted. The proportion of telework was higher for men (21%) than for women (15%) (Table 1).

**Table 1.**
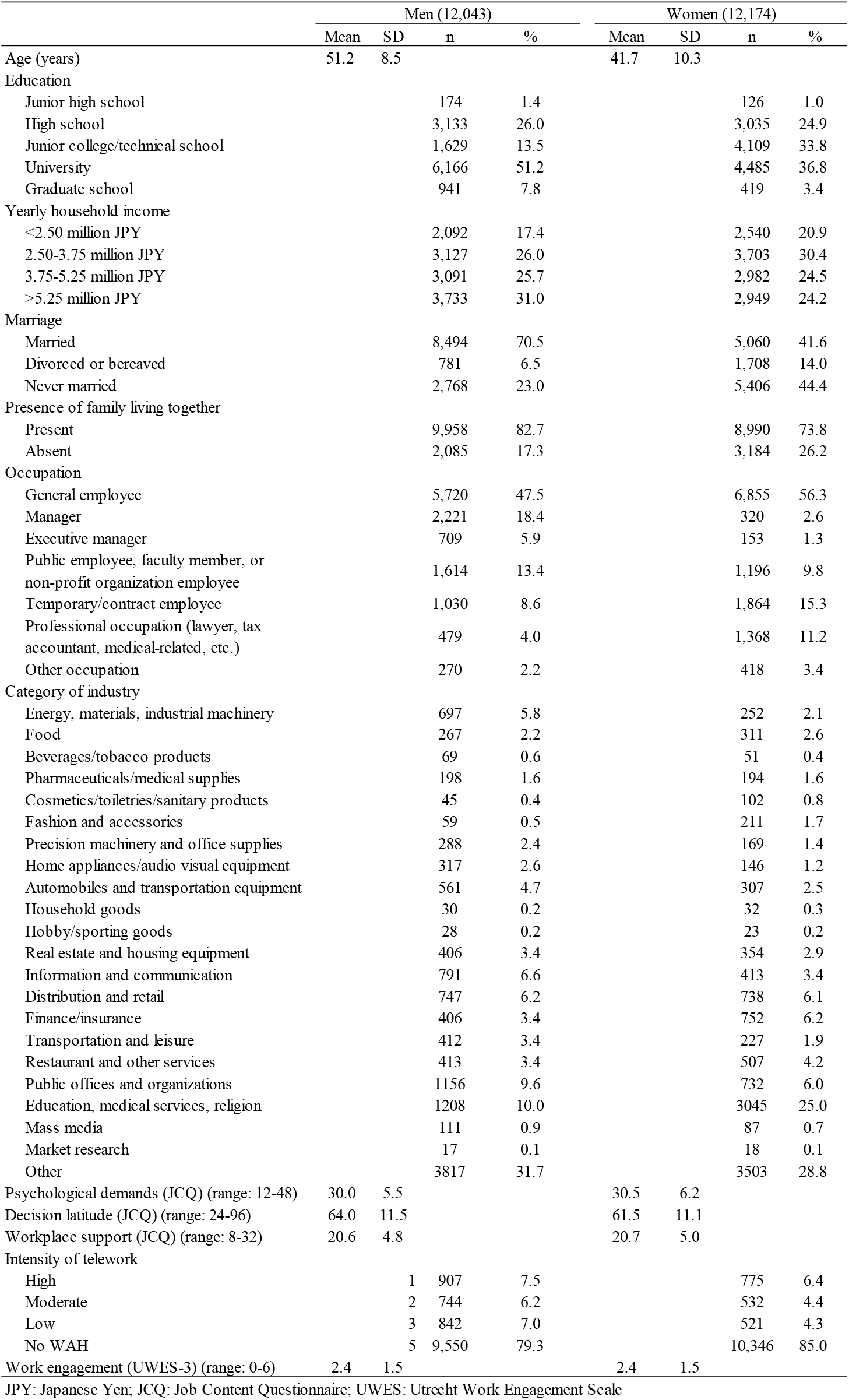
Demographic characteristics, intensity of home-based telework, and work engagement among participants in this study by sex. (n=24,217)

Among all subjects (men and women), all intensity categories of telework were significantly associated with work engagement adjusted for age and sex (Model 1). After adjusting for demographics, including socioeconomic factors, occupation and industry (Model 3), all intensity categories of telework were also associated with work engagement (Table 2). This association remained significant after additionally adjusting for psychological demands, decision latitude, and workplace support for low- and moderate-intensity telework, although not for high-intensity telework (Model 4).

**Table 2.** Association between intensity of home-based telework and work engagement. (12,043 men and 12,174 women)

|  | Model 1 |  |  |  | Model 2 |  |  |  | Model 3 |  |  |  | Model 4 |  |  |  |
| --- | --- | --- | --- | --- | --- | --- | --- | --- | --- | --- | --- | --- | --- | --- | --- | --- |
|  | Coefficient | SE | p value | p for trend | Coefficient | SE | p value | p for trend | Coefficient | SE | p value | p for trend | Coefficient | SE | p value | p for trend |
| Overall (reference = no telework) |  |  |  | <0.001 |  |  |  | <0.001 |  |  |  | <0.001 |  |  |  | <0.001 |
| High-intensity telework | 0.241 | 0.039 | <0.001 |  | 0.232 | 0.038 | <0.001 |  | 0.160 | 0.040 | <0.001 |  | 0.066 | 0.037 | 0.074 |  |
| Moderate-intensity telework | 0.304 | 0.044 | <0.001 |  | 0.242 | 0.044 | <0.001 |  | 0.237 | 0.044 | <0.001 |  | 0.122 | 0.041 | 0.003 |  |
| Low-intensity telework | 0.403 | 0.042 | <0.001 |  | 0.332 | 0.042 | <0.001 |  | 0.317 | 0.042 | <0.001 |  | 0.204 | 0.039 | <0.001 |  |
| Men (reference = no telework) |  |  |  | <0.001 |  |  |  | <0.001 |  |  |  | <0.001 |  |  |  | 0.006 |
| High-intensity telework | 0.214 | 0.052 | <0.001 |  | 0.210 | 0.052 | <0.001 |  | 0.140 | 0.054 | 0.009 |  | 0.058 | 0.051 | 0.250 |  |
| Moderate-intensity telework | 0.326 | 0.057 | <0.001 |  | 0.254 | 0.057 | <0.001 |  | 0.228 | 0.057 | <0.001 |  | 0.109 | 0.053 | 0.041 |  |
| Low-intensity telework | 0.451 | 0.054 | <0.001 |  | 0.360 | 0.054 | <0.001 |  | 0.337 | 0.054 | <0.001 |  | 0.238 | 0.050 | <0.001 |  |
| Women (reference = no telework) |  |  |  | <0.001 |  |  |  | <0.001 |  |  |  | <0.001 |  |  |  | 0.012 |
| High-intensity telework | 0.274 | 0.057 | <0.001 |  | 0.256 | 0.057 | <0.001 |  | 0.179 | 0.059 | 0.002 |  | 0.064 | 0.054 | 0.236 |  |
| Moderate-intensity telework | 0.277 | 0.068 | <0.001 |  | 0.225 | 0.068 | 0.001 |  | 0.257 | 0.068 | <0.001 |  | 0.153 | 0.062 | 0.014 |  |
| Low-intensity telework | 0.329 | 0.068 | <0.001 |  | 0.280 | 0.068 | <0.001 |  | 0.280 | 0.068 | <0.001 |  | 0.157 | 0.062 | 0.012 |  |

Model 1: adjusted for sex, age
Model 2: Model 1 and additionally adjusted for income, marriage, and presence of family living together
Model 3: Model 2 and additionally adjusted for occupation and industry
Model 4: Model 3 and additionally adjusted for psychological demands, decision latitude and workplace support
SE: standard error
High-intensity telework: working at home four or more days per week; Moderate-intensity telework: working at home two to three days per week; Low-intensity telework: working at home once per month to one day per week

In the sex-stratified analysis, after adjusting for demographic factors, including socioeconomic factors and occupation and industry (Model 3), all intensity categories of telework were significantly associated with work engagement. This association remained significant after additionally adjusting for psychological demands, decision latitude, and workplace support for low- and moderate-intensity telework, although not for high-intensity telework for both sexes (Model 4).

## Discussion

The present study revealed an association between home-based telework and work engagement, although the trend differed depending on intensity of telework. High-intensity telework (≥4 days per week) was not associated with high work engagement, while low-intensity telework (once per month to once per week) and moderate-intensity telework (2-3 days per week) had high work engagement for both men and women.

A previous study revealed an indirect relationship between extent of telecommuting and work engagement via social support, but no direct relationship between telecommuting and work engagement was noted.^6^ The authors analyzed the relationship between work engagement and telecommuting using seven levels of intensity in a week as a continuous variable. In the present study, we classified telework into categories based on intensity and conducted analyses by each category, resulting in the demonstration that telework with low-to-moderate intensity was associated with the possibility of increasing work engagement. Telework with an appropriate frequency may increase work engagement. Autonomy has been shown to play an important role in the relationship between telecommunication intensity and job satisfaction.^10^ The key to making telework function more productively is to adopt a management style suitable to telework based on trust and management between supervisor and colleagues and among individual colleagues.^20^ If workers are able to work autonomously and the company is able to provide a suitable working environment for them, workers engaged in high-intensity telework may still be able to maintain high work engagement. A previous study in Japan revealed an increase in labor productivity with a suitable number of teleworking hours; however, when teleworking hours were too long, labor productivity was reduced^13^, suggesting that telework may have negative health effects if overloaded.

Our analysis also showed similar results to a previous study^6^ when the workplace environment factors of psychological demands, decision latitude, and workplace support were added as covariates, suggesting that these factors strongly influence the relationship between telework and work engagement. Determining the ideal intensity of telework may be difficult for workers during the COVID-19 pandemic, depending on their company’s infection prevention measures. In particular, telework has been shown to reduce workplace support from supervisors and colleagues, so the implementation of support measures using Information and Communication Technology tools, such as web conferences, should be considered.^6,20^

In the present study, workers with low- and moderate-intensity telework showed higher work engagement than those with no telework. Workers who are raising children spend more time engaged in housework than those without children, so flexibility of time is important to these workers.^12^ However, telework makes switching between work time and personal time difficult. Stress responses experienced by individuals have been found to propagate across domains, from one area of life within an individual (e.g. work) to another (e.g. family life); this phenomenon is referred to as “crossover”.^21^ Previous studies have shown that working at home increases work stress, and work-family conflict mediates the effect.^22^ Further, the degree of effect was stronger for women than for men. The present study did not take into account work-family conflict in its analysis, so further studies on this point are required. Studies in the United States have shown that home-based telework does not reduce work-family conflict and may actually increase working hours.^23^ Occupational health practitioners need to pay attention to this point when assessing the health impact of telecommuting on workers.

The present study is the first to show an association between the intensity of telework and work engagement under the COVID-19 pandemic. However, several limitations associated with our study warrant mention. First, as the present study was conducted through the Internet, the extent to which the results may be generalized is unclear. However, to reduce bias as much as possible, we sampled the target population according to region, job type, and prefecture based on the infection incidence rate. Second, while work-family conflict may have influenced our findings, we did not enquire about such conflict in this study. However, we did adjust for marital status and the presence of family living together, which may have helped compensate for this lack of data. Third, because this was a cross-sectional study, the causal relationship between the intensity of telework and work engagement is unclear. Concerns have been raised about the existence of reverse causality, such as not choosing telework because the task lowers work engagement. Research has been conducted to index the ease of telecommuting (feasibility of telework) based on job characteristics.^24^ In the present study, we adjusted for occupation and industry, which may have eliminated some of the effects of the feasibility of telework.

In conclusion, low- and moderate-intensity telework (once per month to three days per week) may have beneficial effects on work engagement. Certain factors associated with high-intensity telework (four or more days per week) may not enhance work engagement; these factors should be clarified, and measures to increase work engagement should be taken.

## Data Availability

The datasets are available from the corresponding author on reasonable request.

## Acknowledgements

The present members of the Study Group are Dr. Yoshihisa Fujino (present chairperson of the study group) and (in alphabetical order by given name) Dr. Akira Ogami, Dr. Arisa Harada, Dr. Ayako Hino, Dr. Chimed-Ochir Odgerel, Dr. Hajime Ando, Dr. Hisashi Eguchi, Dr. Kazunori Ikegami, Dr. Keiji Muramatsu, Dr. Koji Mori, Dr. Kyoko Kitagawa, Dr. Masako Nagata, Dr. Mayumi Tsuji, Ms. Ning Liu, Dr. Rie Tanaka, Dr. Ryutaro Matsugaki, Dr. Seiichiro Tateishi, Dr. Shinya Matsuda, Dr. Tomohiro Ishimaru, Dr. Tomohisa Nagata, Dr. Yosuke Mafune. All members are affiliated with the University of Occupational and Environmental Health, Japan.

